# Genome-wide association study suggests a critical contribution of the adaptive immune system to chronic post-surgical pain

**DOI:** 10.1101/2023.01.24.23284520

**Authors:** Marc Parisien, Roel R.I. van Reij, Samar Khoury, Eda Koseli, Mohamad Karaky, Nynke J. van den Hoogen, Garrie Peng, Massimo Allegri, Manuela de Gregori, Jacques E. Chelly, Barbara A. Rakel, Eske K. Aasvang, Henrik Kehlet, Wolfgang F.F.A. Buhre, Camron D. Bryant, M. Imad Damaj, Irah L. King, Jeffrey S. Mogil, Elbert A.J. Joosten, Luda Diatchenko

## Abstract

Chronic post-surgical pain affects a large proportion of people undergoing surgery, delaying recovery time and worsening quality of life. Although many environmental variables have been established as risk factors, less is known about genetic risk. To uncover genetic risk factors we performed genome-wide association studies in post-surgical cohorts of five surgery types— hysterectomy, mastectomy, abdominal, hernia, and knee— totaling 1350 individuals. Genetic associations between post-surgical chronic pain levels on a numeric rating scale (NRS) and additive genetic effects at common SNPs were evaluated. We observed genome-wide significant hits in almost all cohorts that displayed significance at the SNP, gene, and pathway levels. The cohorts were then combined via a GWAS meta-analysis framework for further analyses. Using partitioned heritability, we found that loci at genes specifically expressed in the immune system carried enriched heritability, especially genes related to B and T cells. The relevance of B cells in particular was then demonstrated in mouse postoperative pain assays. Taken altogether, our results suggest a role for the adaptive immune system in chronic post-surgical pain.

## INTRODUCTION

Chronic post-surgical pain (CPSP) is a debilitating condition affecting between 5–85% of patients undergoing surgery, varying with the type of surgery.[1] Chronic pain is defined as pain persisting for longer than 3 months, and may include both nociceptive (inflammatory) and neuropathic components.[1] CPSP has a large negative impact on the quality of life of the affected patients.[1-3]

Despite being a prevalent condition, research on CPSP has been sparse, with the consequence that the pathophysiology of CPSP is only partially understood. Clinical (e.g., type of surgery, surgical skill), demographic, and psychosocial risk factors have been identified to predispose for CPSP,[2, 4] but these only partially explain the observed variance. At present, a basic understanding of the genetic risk factors for CPSP as well as its molecular etiology is still lacking.[2, 4] Studies in other chronic pain syndromes have shown that chronic pain is moderately heritable, with estimates ranging from 30–70% with a median of 45%.[5-9] Candidate gene-based studies on CPSP have provided conflicting evidence.[10-13] Nonetheless, CPSP has recently been introduced as a distinct disease entity in the International Classification of Disease v.11 (ICD-11), which should raise awareness and increase interest in CPSP research.[1]

One way forward to decipher the complex etiology of CPSP is via genome-wide association studies (GWAS), which provide a hypothesis-free tool for identification of genetic contributors.[5] Thus far, one small GWAS of CPSP has been conducted, but no genome-wide significant risk locus was identified.[14] We performed GWAS of CPSP in six independent cohorts covering five different surgical types—hernia repair, hysterectomy, abdominal surgery, knee surgery, and mastectomy—in adults of both sexes. This was followed by a meta-analysis of the independent cohorts, as well as functional analyses. We found that the adaptive immune system rather than the nervous system appears to be the main origin of CPSP. Mouse studies provided positive evidence for the important involvement of B cells in particular.

## METHODS

### Patient Enrolment

Protocols for this study were reviewed and approved by the local Research Ethics Committees relevant to each independent cohort, with details following. All participants gave written informed consent. The HYS cohort was registered at the Dutch trial registry under the number NTR2702 (https://trialsearch.who.int/Trial2.aspx?TrialID=NTR2702), with approval from the Institutional Ethics Comittee of the Maastricht University Medical Center+. Cohorts registered at the Clinical Trials registry (https://clinicaltrials.gov/) were: the ABKNEE cohort under numbers NCT02002663 and NCT01989351, with the approval from the Institutional Review Board (IRB) of the Fondazione Policlinico San Matteo; the TANK cohort under NCT01364870, with the approval from the IRB of the University of Iowa Human Subjects Office; the HRN cohort under NCT00649142, with the approval from the IRB of The Lundbeck Foundation (Scherfigsvej 7, DK-2100 Copenhagen Ø); and the TKR cohort under NCT01557751, with the approval from the IRB of the University of Pittsburg. The PMPS cohort was not registered in a public trial registry database as it was an observational study, but conducted with the approval from the IRB of McGill University’s Faculty of Medecine.

Elaborate descriptions of patient recruitment, sample, and data collection protocols for the cohorts have been published previously (Supplementary Table 1).[14-20] In brief, a multi-center cohort study was conducted in four hospitals in the Netherlands (hysterectomies, HYS), three hospitals in Italy (abdominal surgeries and knee replacements, ABKNEE), one hospital in Pittsburgh, USA (mastectomies, PMPS; and total knee replacements, TKR), one hospital in Denmark (hernia repair, HRN), and two hospitals in Iowa, USA (total knee arthroplasties, TANK).

### Genotyping

In the PMPS cohort the DNA samples were derived from lymph node tissue, blood, or saliva; in the rest of the cohorts DNA was purified from blood samples. HYS and ABKNEE cohorts were genotyped at the Department of Genomics at the Life and Brain Center, University of Bonn, using the Illumina PsychArray (Infinium PsychArray-24 v1.2 Bead Chip, Illumina Inc., USA). The PMPS, HRN, TKR, and TANK cohorts were genotyped using the UK Biobank Axiom platform at the Genome Center at McGill University. Genotypes were called using BeadStudio (Genome Studio v2011.1, Illumina Inc, USA). A standard GWAS cleaning protocol[21] was applied using PLINK version 1.9[22, 23] in each cohort independently.

Genotype imputation for all cohorts except PMPS was performed using the stepwise imputation approach implemented in Minimac3 [24] and Eagle2 v2.3,[25] using default parameter settings and the European HRC reference panel version r1.1.[24-26]. The PMPS cohort was first pre-phased with SHAPEIT,[27] followed by IMPUTE2 for imputation,[28] using the 1,000 Genomes Project phase III reference panel.

The sample quality control (QC) procedures consisted of discarding individuals prior to statistical analysis based on five criteria (Supplementary Table 2): 1. Missing pain phenotype; 2. SNP missingness rate > 5%; 3. Heterozygosity rate deviating by more than three standard deviations indicating peculiar heterozygosity; 4. Identity by descent (IBD) π > 0.185 indicating relatedness to an individual already selected with a better genotyping rate (up to 3^rd^ degree relative); 5. Multidimensional scaling (MDS) of more than five standard deviations. A total of 1350 participants of Caucasian ancestry passed the sample QC.

SNP-based QC consisted of discarding variants based on several criteria (Supplementary Table 3): 1. Missingness rate > 5%; 2. Deviation from Hardy-Weinberg equilibrium P < 1×10^−6^; 3. Effective minor allele count < 10, where the effective minor allele count defined as: imputation info score * 2 * number of individuals * minor allele frequency. In all, between five to eight million markers were available after QC.

### Genome-Wide Association Studies

The cohorts were re-organized in a way defined by surgery type (Supplementary Table 4). This way, the ABKNEE cohort was split in two; 90 subjects with abdominal surgery while 71 with knee arthroplasty. Likewise, all three knee surgery cohorts were grouped together under “knee” with now 303 total individuals.

Genome-wide association analyses were carried out using PLINK (v1.9).[22, 23] We assumed an additive inheritance model on non-rare variants (minimum of 10 effective minor allele counts). The primary outcome, pain, was corrected for self-declared sex as well as the top five genetic principal components. A total of seven GWAS were conducted: 1. HYS; 2. Abdominal surgeries of ABKNEE; 3. Knee surgeries of ABKNEE; 4. PMPS; 5. HRN; 6. TKR; and 7. TANK. Then, a meta-analysis was conducted using METAL[29] to combine all three knee surgery cohorts into one. Finally, we combined all seven GWAS using METAL again. In METAL, we enabled the fixed-effect, standard-error-weighted scheme “SCHEME STDERR”, as well as the “GENOMICCONTROL ON” option which will remove any inflation from input GWAS test statistics. Retained variants appeared in at least two of the merged cohorts and displayed allele frequency heterogeneity P-value > 10^−4^. Overall, we obtained GWAS summary results for each of the five surgery types, and one for all cohorts combined.

The primary outcome measured in the HYS and PMPS cohorts was the highest surgery-related pain score measured on a numeric rating scale (NRS), recorded at rest 8-9 weeks post-surgery.[14, 15] The primary outcome measured in the ABKNEE cohort was the average surgery-related pain score measured on the NRS, recorded at rest during the last week, three months post-surgery.[14] The primary outcome in the TANK and TKR cohorts were assessed using the Brief Pain Inventory (BPI) “worst pain” at six months after surgery. The primary outcome in the HRN cohort was assessed by NRS at six months after surgery. All primary outcome measures were assessed on a continuous scale. About 21.5% of individuals displayed pain scores ≥ 4: hysterectomy 10%, abdominal 1%, mastectomy 30%, hernia 15%, and knee 27%.

### Functional characterization of findings

MAGMA v1.07b was used to obtain gene- and pathway-level summary statistics from SNP-level ones, from European population reference data.[30] Gene definitions were those based on NCBI’s human genome build 37, distributed alongside MAGMA (https://ctg.cncr.nl/software/magma), whereas pathways were those from Gene Ontology’s biological processes,[31] downloaded December 2020 from http://download.baderlab.org/EM_Genesets/. For gene-based analyses, MAGMA was run with the ‘--gene-model multi=snp-wise’ option, which estimates gene-level statistics based on mean SNPs effects but combined with the top 1 SNP effect. This way, the gene-based Manhattan plot is more congruent with the SNP-based one, which is biologically more intuitive.

The percent variance explained for the CPSP phenotypes by genetic variability were estimated using GCTA, and evaluated directly on the quantitative NRS scale rather than assuming a 10% prevalence if stratified by dichotomous pain (≥4) versus no pain (<4).[1, 32, 33]

Tissue-specific partitioned heritability was determined with LDSC.[34, 35] The sources for tissues and cell type gene expression was from the dataset published by Benita et al.,[36] a gene expression database comprised of 126 human samples, including several from the central nervous system, the peripheral nervous system, muscles, and various immune cell types, downloaded from http://xavierlab2.mgh.harvard.edu/EnrichmentProfiler/Datasets.html. For each cell line or tissue, we extracted the top 1,000 most-specifically expressed (i.e., enriched) genes and used these for partitioning heritability. For such a large number of genes (as suggested in LDSC’s work), the goal was to establish an overall transcriptome signature rather than trying to highlight a few dozen uniquely expressed genes. We then replicated our results in the dataset published by Fehrmann et al.,[37] and in the multi-tissue chromatin dataset published by Trynka et al.,[38] available at the Broad Institute: https://data.broadinstitute.org/alkesgroup/LDSCORE/.

### Mouse Studies

#### Mice

In order to validate the results obtained by the genetics analysis, we examined pain behavior in two mouse assays of postoperative pain. Wildtype (C57BL/6) and *Rag1* null mutant (*Rag1*^-/-^; B6.129S7-*Rag1*^tm1Mom^/J) mice of both sexes were purchased from The Jackson Laboratory (Bar Harbor, ME). Mice were housed same-sex in standard shoebox cages (with enrichment) with 2-4 mice per cage, maintained in a temperature controlled (20 ± 1 °C) environment (12:12 h light/dark cycle; lights on at 07:00 h), and received rodent chow (Envigo Teklad) and filtered municipal tap water *ad libitum*. Mice were acclimated for at least one week to the vivarium before any testing occurred. All experiments were approved by local animal care and use committees at McGill University or Virginia Commonwealth University (details below).

#### Plantar incision

All mice received plantar incision surgery in the right hind paw as originally described by Brennan et al.[39] Mice were anesthetized with 5% isoflurane at 2 L/min delivered into a sealed induction chamber then maintained with 2.5% isoflurane at 500 mL/min with a nose cone. A 5-mm longitudinal incision was made in the plantar paw, the flexor digitorum brevis muscle was elevated, then a longitudinal incision was made through it. The wound was closed by two sutures in the skin and covered by Bacitracin zinc ointment antibiotic (Polysporin). All surgeries were performed by the same surgeon (MK). Once recovered from anesthesia, mice were returned to their cages. The experiments were approved by McGill University’s Animal Care and Use Committee, and conformed to the Canadian Council on Animal Care guidelines.

#### Laparotomy

Laparotomy surgeries were performed as described by Oliver et al. with slight modifications.[40] Mice were anesthetized with 5.0% isoflurane and maintained using a constant stream of 2.5% isoflurane in oxygen using a face mask and vaporizer. Abdominal hair was clipped with battery-powered clippers and the surgical site was disinfected with beta-iodine. All surgeries were performed by the same surgeon (EU). A 1.0-to 1.5-cm midline abdominal incision was made through the skin and then extended through the linea alba. To mimic visceral manipulation associated with various surgeries, a sterile cotton swab was inserted and moved throughout the abdomen for 30 s. The abdominal muscle layers and skin was closed by using 5-0 nylon suture in a discontinuous pattern. Once recovered from anesthesia, mice were returned to their cages. Every surgical group’s corresponding sham group received the same duration of isoflurane at the same time and their hair was clipped and surgical site disinfected but no incisions were made. The experiments were approved by The Institutional Animal Care and Use Committee of Virginia Commonwealth University.

#### Behavioral tests

In the hind-paw incision experiments, mechanical sensitivity measurements were made before and after the surgery at day 1, 4, 7, and then weekly up to day 84 post-surgery. To do so, mice were placed individually in transparent Plexiglas cubicles (5 cm by 8.5 cm by 6 cm) for 1 hour to be acclimated, then withdrawal responses to mechanical stimuli were determined using calibrated von Frey filaments (force range: 0.008 to 1.4 g; Touch Test Sensory Evaluator Kit, Stoelting) applied to the plantar surface of each hind paw. The withdrawal thresholds were calculated using the up-down method of Dixon.[41] In the laparotomy experiments, abdominal sensitivity was assessed with a scoring system that was adapted from Schiene et al.,[42] and consisted of a scale of 0–3 for the withdrawal reactions following six abdominal stimulations with a 0.4 g von Frey filament (1 stimulation every 2 min), where a score of 0 indicated no response; 1 indicated lifting of the abdomen, licking or movement; 2 indicated hind-paw extension or flinching, slight jumping or strong licking; and 3 indicated strong jumping or running inside the chamber. The pain score was the sum of scores for all stimulations, out of a total of 18. Mice were tested at baseline and at 1, 4, 7 14, 21 and 28 days post-surgery.

#### Immune cell rescue experiment

Separate groups of mice were injected in the tail vein with saline or isolated B and/or T cells. Two weeks later, mice were tested for mechanical sensitivity before 1, 3, 7 and 10 days post-hind paw incision surgery as described above.

### Statistical Analyses

For each statistical test, the threshold for significance is indicated, as well as the methods used for correction for multiple testing (Bonferroni, false discovery rate, etc.).

For GWAS, linear regression was used to assess the association between allelic dosage and pain intensity.[43, 44] We complimented PLINK’s linear regression model with other models tested with the R statistical package, including Poisson, quasi-Poisson, negative binomial, etc. and found that the linear regression provided the most flat, less skewed distribution of association P-values (data not shown), which ideally should be uniformly distributed (given a fair null model of association, e.g., linear regression of the phenotype to the additive genetic inheritance burden model).

Mouse data were analyzed using GraphPad Prism, v.7.04 (GraphPad Software, Inc., La Jolla, CA). Normality and homoscedasticity of all data sets were confirmed by using the Shapiro–Wilk and Levene tests, respectively. Data were analyzed using repeated measures ANOVA, followed by posthoc testing as appropriate. A criterion α =0.05 was adopted for statistical significance.

## RESULTS

### Genome-Wide Association Studies

Following QC, a total of 1350 individuals with simultaneous genotype and phenotype information enabled association studies at the genome-wide scale, first on a per-surgery-type basis, followed by a meta-analysis of all cohorts (Table 1). Genome-wide association results are displayed in Manhattan plots (Fig. 1), for SNP-wise summary results (green), gene-wise ones (blue), and pathway-wise ones (purple). There, we observed many hits reaching statistical significance (Bonferroni’s genome-wide 5×10^−8^ threshold for SNPs, FDR 20% for genes and pathways) (Supplementary Table 5). Significant SNP hits were grouped in loci in which the lead SNP was highlighted along genes residing immediately before, within, and immediately after these hits (Table 2). QQ plots indicated statistical significance at SNP, gene and pathway levels guarding against false discovery rates of up to 20% (Fig. 1; Supplementary Table 5).

**Table 1.** Summary of GWAS results per surgery type. Genome-wide significance threshold set at 5×10^−8^, while nominal significance at 5×10^−7^.

| Surgery | GWS | Loci | NOMS | $\lambda$ GC | PVE (%) | $P_{PVE}$ |
| --- | --- | --- | --- | --- | --- | --- |
| hysterectomy | 24 | 12 | 107 | 1.03 | 99 | 0.07 |
| abdominal | 3 | 2 | 21 | 1.03 | 99 | 0.33 |
| mastectomy | 0 | 0 | 0 | 1.02 | 83 | 0.14 |
| hernia | 44 | 6 | 69 | 1.02 | 64 | 0.36 |
| knee | 1 | 1 | 1 | 1.02 | 12 | 0.41 |
| meta-analysis | 5 | 3 | 22 | 1.00 | 39 | 0.02 |
| total | 77 | 24 | 220 |  |  |  |

Abbreviations are: genome-wide significance, GWS; nominal significance, NOMS; genomic control's lambda, $\lambda$ GC; percent variance explained of the phenotype by genetics, PVE; associated P-value, $P_{PVE}$ .

**Table 2.**
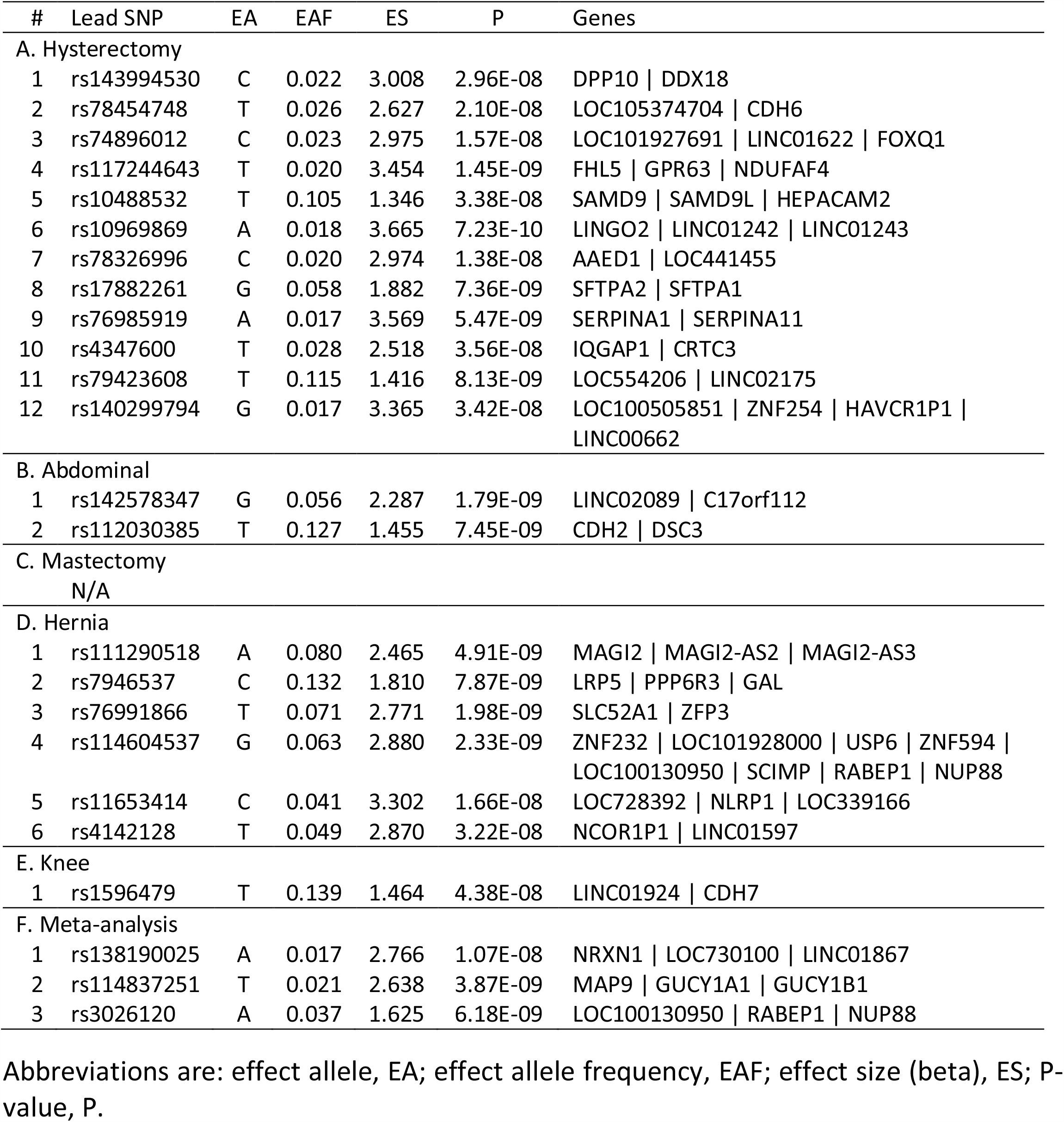
Summary of genome-wide significant hits. SNP hits are grouped by surgeries (A to E) and by overall meta-analysis (F). Only the lead SNP per locus is shown.

**Figure 1.**
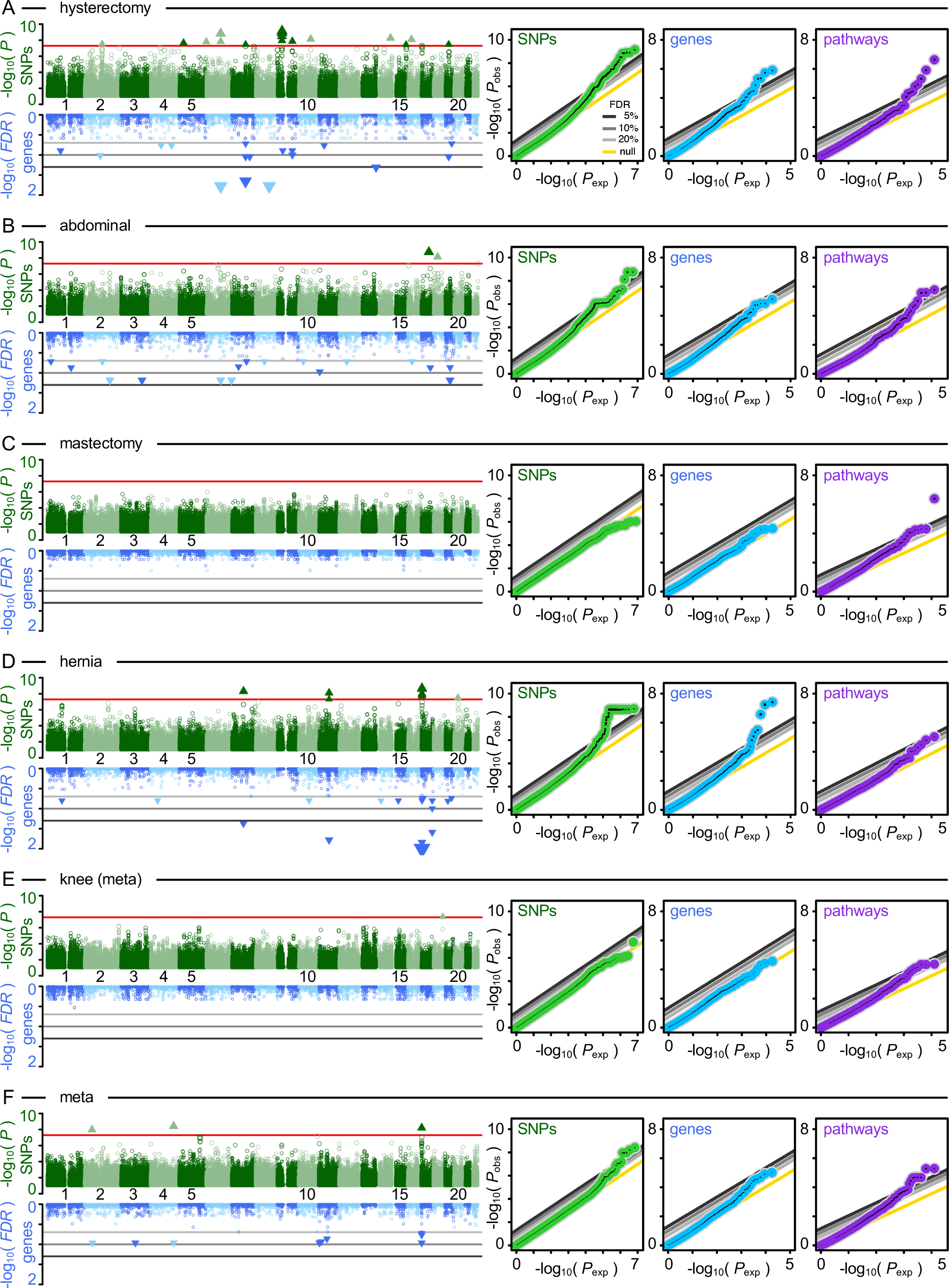
Association summary statistics by surgery types. Summary statistics are shown using: Manhattan plots (left) for SNP-level (top, green) and gene-level (bottom, blue) summaries, as well as QQ plots (right) for SNP-level (green), gene-level (blue), and pathway-level (purple) summaries. Manhattan plots use dark color hues for odd chromosomes while light color hues for even ones. SNP-level genome-wide significance indicated using red horizontal bars, while gene-level genome-wide significance is indicated using various false discovery rate (FDR) thresholds ranging from 5 (black) to 10% (dark grey) to 20% (light grey). QQ plots track observed P-values (Pobs) as a function of expected ones (Pexp). SNP-level statistics obtained with PLINK or METAL (meta-analysis), while gene-level statistics obtained with MAGMA. Surgeries are: **(A)** hysterectomy, **(B)** abdominal, **(C)** mastectomy, **(D)** hernia, and **(E)** knee, from a meta-analysis. **(F)** meta-analysis of all cohorts combined.

Seventy-seven genome-wide significant SNP hits were uncovered scattered across 24 loci, 244 genes, and 64 pathways (Supplementary Table 5). Only the mastectomy GWAS didn’t yield any nominally significant SNP hits (P<10^−7^). Among the 24 significant genetic loci, one locus spanning the genes LOC100130950, RABEP1, and NUP88 was found significant in both the hernia GWAS as well as in the meta-analysis of all cohorts. All point estimates for percent phenotypic variance explained by genetics, also known as narrow-sense heritability, were found to be non-significant (P>0.05), except for the overall meta-analysis with 39% variance of the CPSP phenotype explained by genetics (P=0.02).

### Tissues or Cell Types with Enriched Heritability

We next tested tissues or cell types whose uniquely expressed genes would carry excess heritability when contrasted against other, non-tissue-specific genes. To do so, we performed partitioned heritability, with partitions based on genes expressed in cell types or tissues pertinent to pain states. The selected gene expression database contained many central nervous tissues, including various brain regions as well as the spinal cord, peripheral nervous system ganglia (in particular, dorsal root and trigeminal), many immune cell types, musculoskeletal, and other tissues and cell types.[36] Overall, 126 tissues and cell types were tested, grouped into eight broad categories (Fig. 2, Supplementary Table 6A). We found enriched heritability at the FDR 20% level in immune cells, more specifically in B and T cell subtypes, and in the CD34+CD38–hematopoietic progenitor stem cell. No significant evidence was found to support neutrophils, macrophages, tissues of the central or peripheral nervous systems, or musculoskeletal tissues. The findings were replicated in another gene expression database, by Fehrmann, itself also containing brain regions as well as immune cell types (Supplementary Table 6B). There, among eight cell and tissue types with enrichment P≤0.05, B and T cell subtypes were significantly enriched. Further support for gene expression in T cells came from the dataset by Trynka et al. with H3K9ac and H3K27ac chromatin markers (P≤0.05), where acetylated H3K9 turns on gene expression, and acetylated H3K27 enhances gene transcription (Supplementary Table 6C). In this dataset, among six cell types with the evidence for enrichment at P<0.05, four were different subsets of T cells. It is unclear which B or T cell subtypes are most critically important in post-surgical pain chronification, and there was no exact correspondence of the blood cell types in the different datasets. Nonetheless, our results overall point to the adaptive immune system rather than to the innate immune system or the nervous system and are consistent with lymphocyte differentiation.

**Figure 2.**
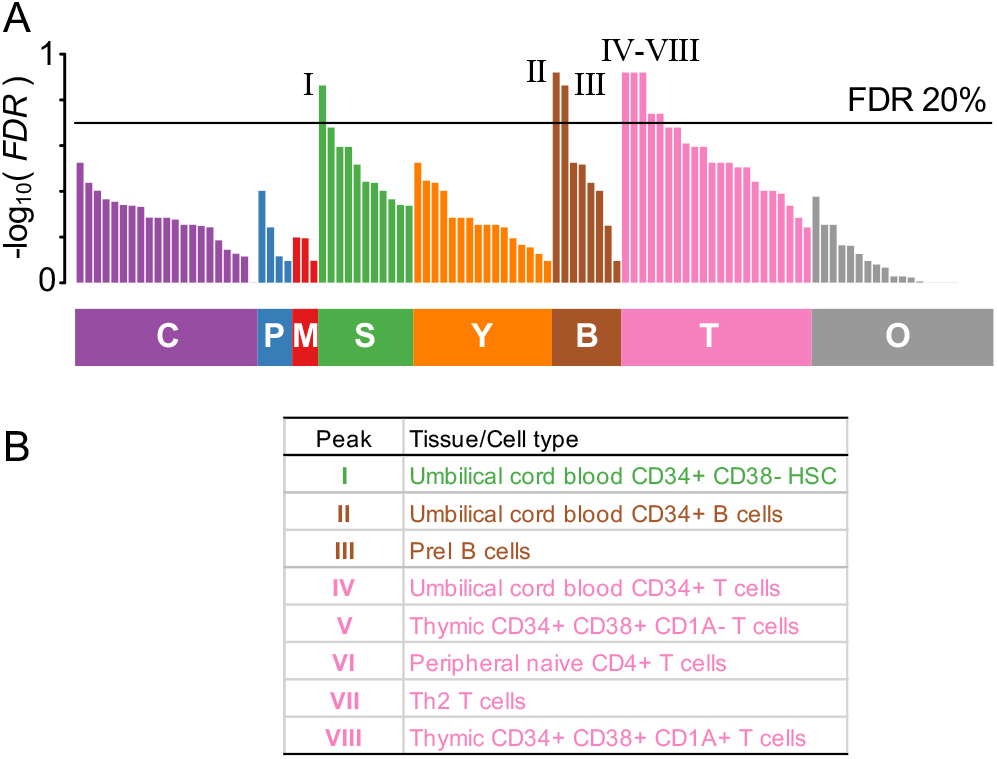
Partitioned heritability in selected tissues in the CPSP meta-analysis. **(A)** Enrichment tracked in 106 tissues or cell lines from Benita et al. Tissues are grouped by: central nervous system (C, purple, n=21), peripheral nervous system (P, blue, n=4), muscle (M, red, n=3), stem cells (S, green, n=11), myeloid cells (Y, orange, n=16), B cells (B, brown, n=8), T cells (T, pink, n=22), and other tissues or cell lines (O, grey, n=21). Shown are FDR-corrected P-values for enrichment. **(B)** Tissues and cell lines significantly enriched at the FDR 20% level, numbered from left to right using Roman numerals as in panel A.

### Mouse Studies

Based on the results from genetic analyses in which partitioned heritability analyses alluded to the plausible involvement of B and T cells, we next proceeded to test the roles for the adaptive immune system in post-surgical pain using mouse assays. To do so, we employed *Rag1*^-/-^ mice, in which a null mutation leads to impaired adaptive immune systems, with mice having no mature B and T lymphocytes.[45] We utilized two well-characterizedpreclinical post-surgical pain assays: plantar incision and laparotomy.[46]

In the plantar incision assay, withdrawal thresholds to evoked mechanical stimulation (using von Frey filaments) aimed at the plantar hind paw before and at multiple time points up to the 84th day after hind-paw incision were measured in C57BL/6 (wildtype) and *Rag1* null mutant (*Rag1*^-/-^) mice lacking T- and B-cells. Repeated measures ANOVA revealed significant interactions of repeated measure with strain and sex (*F*_13,221_ = 2.1, *p*=0.015). As seen in Supplementary Figure 1, however, both sexes clearly displayed both robust allodynia and a large genotype effect. Collapsing across sex, the repeated measure by strain interaction was highly significant (*F*_13,247_ = 2.3, *p*=0.007). As shown in Fig. 3A, whereas the mechanical allodynia produced by the incision was fully resolved in wildtype mice by 5 days post-surgery, an exacerbated and greatly prolonged allodynia (with a statistically significant strain difference, corrected by multiple comparisons, up to 42 days post-surgery) was observed in *Rag1*^-/-^ mice. There was no main effect of repeated measure (*F*_13,247_ = 1.1, *p*=0.29) or repeated measure by strain interaction on the contralateral hind paw (*F*_13,247_ = 1.7, *p*=0.07) (not shown).

**Figure 3.**
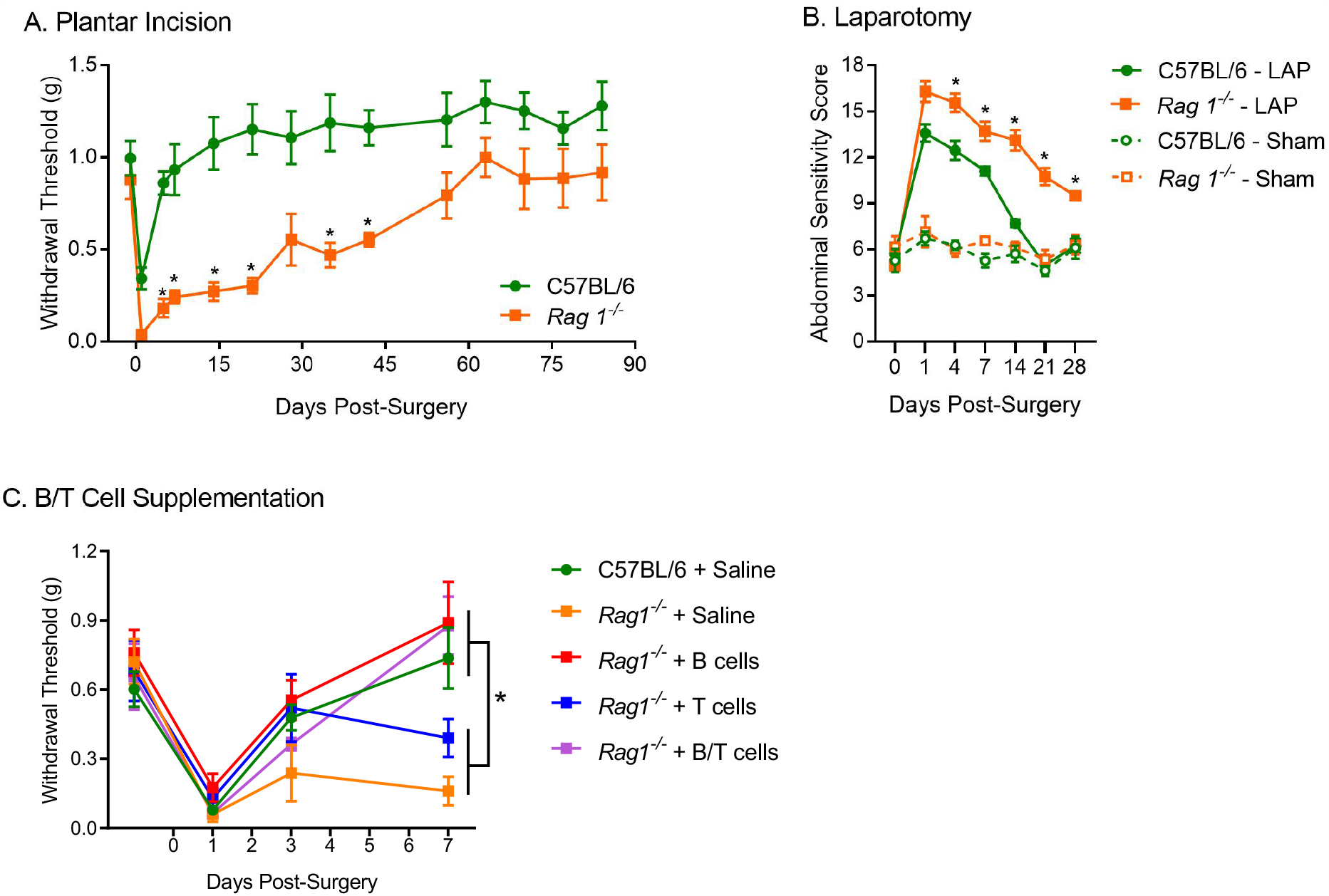
Rag1^-/-^ mice lacking T and B cells display increased and prolonged postsurgical pain. (**A**) Mechanical pain thresholds before and after plantar incision in the ipsilateral hind paw in C57BL/6 (wildtype; *n*=11) and *Rag1*^-/-^ null mutant mice (*n*=10). (**B**) Abdominal sensitivity scores (see Methods) before and after laparotomy (LAP) or sham surgery in C57BL/6 and *Rag1*^-/-^ mice (n=8 per surgery per genotype), performed in a different laboratory. (**C**) Supplementation of B cells resolves pain hypersensitivity in *Rag1*^-/^ mice. Mice received saline, or tail vein injection of T cells, B cells, or both (*n*=5 per group). Symbols in all graphs represent mean ± SEM. \**p*<0.05 compared to wildtype (**A, B**) or as indicated (**C**).

A similar pattern of responses was observed in the laparotomy assay, No main effects of, or interactions with sex were observed, so data were collapsed across sex. A two-between (genotype, surgery), one-within repeated measures ANOVA revealed significant main effects and interactions, including a genotype x surgery x repeated measures interaction (*F*_6,49_ = 6.4, *p*<0.001). Pain hypersensitivity in this assay was observed only in mice given laparotomy, and increased and prolonged in duration in the *Rag1*^-/-^ mutant mice (Fig. 3B). This experiment improves both external validity (generalizability across another surgery type) and internal validity (replication in an independent laboratory) to the conclusion that T and/or B cells are relevant to post-surgical pain in mice.

Since *Rag1*^-/-^ mice are deficient in both T- and B-cells, a new study was performed in male mice in which these cells were individually or jointly replaced (compared to mice receiving saline). ANOVA revealed a significant repeated measures by strain interaction (*F*_4,76_ = 2.7, *p*=0.002). As shown in Fig. 3C, the prolonged duration of mechanical allodynia in untreated *Rag1*^-/-^ mice compared to wild-type mice was observed again. T cell treatment did not significantly affect the time course of mechanical allodynia in *Rag1*^-/-^ mice. However, B cell supplementation, and B plus T cell supplementation, were both able to resolve allodynia at Day 7 and 10 post-surgery (all *p*’s<0.05 compared to *Rag1*^-/-^ plus saline).

## DISCUSSION

The aim of this study was to identify genetic variants associated with CPSP severity in the general surgical population. We employed a GWAS approach in six independent cohorts grouped into five surgical types to interrogate the common variants associated with CPSP severity. Although this was a GWAS study of several relatively small-scale studies, we were able to identify the first genome-wide significant hits associated with CPSP severity in the majority of the surgeries and the meta-analysis. This study will be the starting point for more large-scale efforts and further detailing the mechanisms of CPSP in general and for specific surgical types.

With this study, we were also able to estimate for the first time the SNP-based heritability of CPSP incidence in a diverse array of surgeries. The estimate of 35% is in line with a recent study on overall chronic pain heritability in the UK Biobank which found an estimate of 38.4%.[47] This further solidifies chronic pain in general and specifically CPSP as a heritable trait. Our GWAS analysis identified a total of 77 genome-wide significant SNPs across 24 loci associated with the severity of CPSP (Table 1). Furthermore, hundreds of genes with top one or aggregated SNP effects as well as biological pathways were found significant at the FDR 20% level (Supplementary Table 5).

After performing GWAS, we did functional analyses of our genetic results to gain insights into biological processes contributing to CPSP. And here our findings were unexpected when we tested tissue-specific partitioned heritability. Previous genome-wide association studies of chronic pain indicated enriched heritability in brain regions, with the DCC gene and the axonal guidance pathway showing the strongest genetic associations.[48-52] These studies pointed to the genetic root for the neuronal origin hypothesis of chronic pain. Interestingly, brain connectivity was found to be a predictor for the transition to chronic pain,[53, 54] while effective treatment of the chronic pain was shown to reverse abnormal brain anatomy and function.[55] Here, rather, we observed an enrichment for heritability in adaptive immune cells, more specifically in B and T cells, while no support for neuronal tissues neither at the central or peripheral nervous systems levels was found. The enrichment pattern seems further suggest differentiation of B and T cells, judged by enrichment for hematopoietic progenitor stem cell. The immune origin of chronic pain is thus highlighted. A possible explanation for this apparent neuro-immune paradox might be in the stronger immune response elicited by the explicit surgery-related injury and following inflammatory responses.[56]

Another explanation for the apparent neuro-immune paradox could be due to the difference in time spans for the assessment of the reported pain. Typically, chronic pain patients have their pain lasting for years, while in our current study the cases were evaluated at the three or six months mark. Possibly, our results point that at the earlier stages of persistent pain, like in case 3-6 months post-surgical pain, the role of the immune system is the most substantial, however, the majority of these cases will resolve over time and for the very long-lasting chronic postsurgical pain, the contribution of the neuronal system is the most critical. Nonetheless, implications of the immune system in chronic pain are already well documented,[57-61] and further supported here via GWAS.

The physiological response to wounding, as occurs in surgical contexts, features a complex yet distinctive profile of coordinated, overlapping, and time dependent events. Traditionally, these events are categorized into three phases that are 1) an APOP-inducing inflammatory reaction, 2) proliferative events that lead to the restoration of affected tissue, and 3) remodelling of the affected tissue.[62, 63]

In all phases, a significant role for T lymphocytes is found. CD4+ T helper cells are attracted by the IFN-gamma secreted by macrophages and contribute to the initial proinflammatory environment, γδT cells promote proliferation of keratinocytes thereby affecting tissue remodeling and regulatory T cells subside the inflammation by secretion of anti-inflammatory mediators.[63, 64] Although the exact role of B lymphocytes within the wound healing process has not yet been identified, it has been shown that CD19 (a key regulator of B cells) controls wound healing through hyaluronan-induced TLR4 signalling process. These LR4-dependent pro-inflammatory signalling induces B cell proliferation and cytokine excretion indicating a substantial role of B lymphocytes.[65] In part, the detrimental effect of prolonged inflammation after injury results from the deregulation and desynchronization of wound-healing events, such as the impediment of the transition towards the proliferation phase in which both B and T cells play critical roles.[64] As corticosteroids are often administed as a standard of care after surgery, their impact on adaptive immunity must be taken into account.[66]

We then validated the results obtained by genetics using B/T-cell-deficient mice. After hind paw or abdominal surgical incision, mice with impaired B and T cells displayed higher peak levels of pain hypersensitivity, but more importantly remained in an allodynic state significantly longer than their wild-type counterparts, indicating a role for B/T cells in the maintenance post-surgical pain (Fig. 3). To identify which immune cell type was directly responsible for this prolonged allodynia, we attempted to rescue the allodynic effect in the *Rag1*^-/-^ mice using isolated B or T cells. The rescue was observed convincingly in the presence of supplemented B cells, thus establishing their critical role (Fig. 3).

There is a number of limitations in this study we would like to mention. 1. Small sample sizes for GWAS; 2. Several types of surgeries, with potentially different pain intensities and chronification rates; 3. The majority of cases were evaluated at the three months post-surgery mark, thus still had chance to resolve their pain by the following six months mark; 4. Our cohorts do not have a balanced male and female representation as the majority of our participants (75%) were women; 5. Absence of *bona fide* replication in independent cohorts; 6. High probability for hospital- and country-specific post-surgical care such as drug treatments, post-surgery care, post-surgical hospital stay length, and diet. 7. Overlap in genes expressed in B and T cells, overlap introduced from requirements of partitioned heritability as the authors suggested as many as 10% of expressed genes, thus veiling the ability to unequivocally distinguish between B and T cells. That being said, the results of the mouse experiments suggest that B cells rather than T cells are the effectors of pain after surgery, displaying a net protective effect.

To conclude, this study found 77 SNPs spread over 24 genomic loci associated with CPSP severity, a debilitating condition affecting between 5 and 85% of patients undergoing surgery and varying with surgery type. Functional analysis strongly suggested a critical contribution of the adaptive immune system in the genetic factors associated with CPSP. More specifically, B cells rather than T cells were able to rescue the prolonged allodynia experienced by immune-compromised mice following incisional surgery at the paw. The insights into the etiology of CPSP severity warrant further studies both clinical and preclinical to further specify the underlying mechanism for CPSP in general and each specific surgical type.

## Data Availability

All data produced in the present study are available upon reasonable request to the authors

## ACKNOWLEDGEMENTS

We thank all participants without which this study wouldn’t have been possible.

## SUPPLEMENTARY TABLE LEGENDS

**Supplementary Table 1.** Study design of chronic post-surgical pain cohorts.

| Surgery | Acronym | Enrolled | Country | Hospitals | Genotyping Array |
| --- | --- | --- | --- | --- | --- |
| hysterectomy | HYS | 345 | Netherlands | A,B,C,D | Illumina PsychArray |
| abdomen & knee | ABKNEE | 190 | Italy | E,F,G | Illumina PsychArray |
| mastectomy | PMPS | 665 | USA | H | Axiom UK Biobank |
| knee replacement | TKR | 100 | USA | H | Axiom Precision Medicine |
| hernia | HRN | 226 | Denmark | I | Axiom Precision Medicine |
| knee arthroplasty | TANK | 180 | USA | J,K | Axiom Precision Medicine |

**Supplementary Table 2.** Samples quality controls.

|  | HYS | ABKNEE | PMPS | TKR | HRN | TANK | Total |
| --- | --- | --- | --- | --- | --- | --- | --- |
| enrolled | 345 | 190 | 665 | 100 | 226 | 180 | 1706 |
| without phenotype | -15 | -27 | -201 | -16 | -41 | -30 | -330 |
| missingness >5% | 0 | 0 | 0 | 0 | 0 | 0 | 0 |
| heterozygosity $\vert Z\vert > 3$ | -3 | -1 | -7 | 0 | -2 | -2 | -15 |
| IBD $\pi_{\text{hat}} > 0.185$ | 0 | 0 | -3 | 0 | 0 | 0 | -3 |
| MDS $\vert Z\vert > 5$ | -2 | -1 | -4 | 0 | -1 | 0 | -8 |
| genotype & phenotype | 325 | 161 | 450 | 84 | 182 | 148 | 1350 |
| females | 325 | 104 | 450 | 53 | 0 | 91 | 1023 |
| % | 100.0 | 64.6 | 100.0 | 63.1 | 0.0 | 61.5 | 75.8 |
| males | 0 | 57 | 0 | 31 | 182 | 57 | 327 |
| % | 0.0 | 35.4 | 0.0 | 36.9 | 100.0 | 38.5 | 24.2 |

Abbreviations are: identity by descent, IBD; multidimensional scaling, MDS; Z-score, Z.

**Supplementary Table 3.** Markers quality controls.

|  | HYS | ABKNEE | PMPS | TKR | HRN | TANK |
| --- | --- | --- | --- | --- | --- | --- |
| imputed | 39,018,346 | 39,018,346 | 81,178,076 | 39,018,346 | 39,018,346 | 39,018,346 |
| missingness >5% | 0 | 0 | -561,428 | 0 | 0 | 0 |
| HWE P <1e-6 | -44 | -37 | -1,011 | -2 | -14 | -12 |
| effective MAC < 10 | -32,120,272 | -33,047,338 | -72,015,576 | -33,955,104 | -32,848,581 | -33,121,718 |
| final | 6,898,030 | 5,970,971 | 8,600,061 | 5,063,240 | 6,169,751 | 5,896,616 |

Abbreviations are: Hardy-Weinberg equilibrium P-value, HWE P; minor allele count, MAC.

**Supplementary Table 4.**
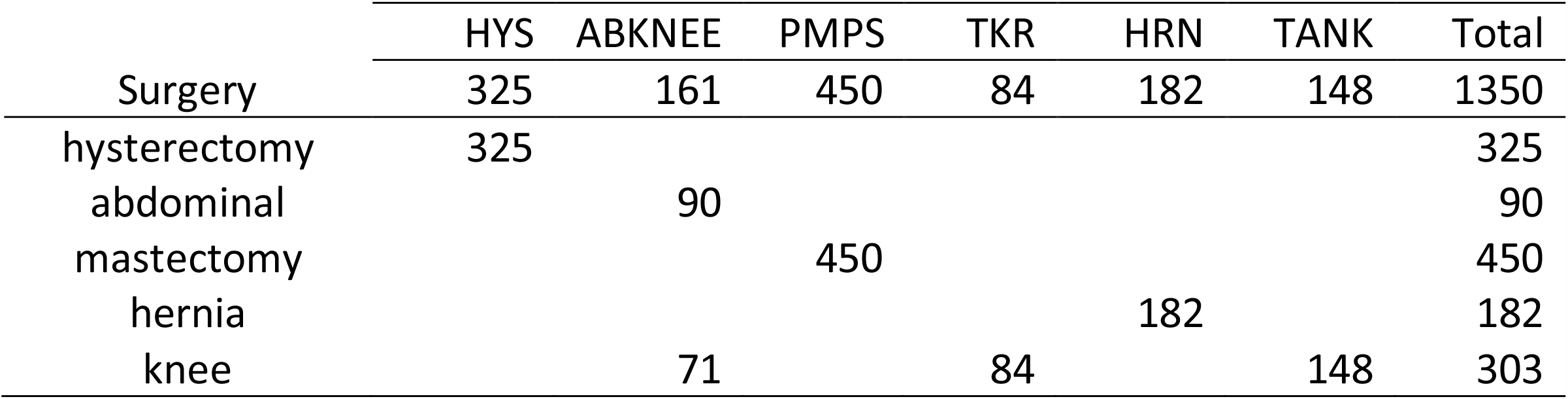
Distribution of individuals per surgery type for GWAS.

|  | HYS | ABKNEE | PMPS | TKR | HRN | TANK | Total |
| --- | --- | --- | --- | --- | --- | --- | --- |
| Surgery | 325 | 161 | 450 | 84 | 182 | 148 | 1350 |
| hysterectomy | 325 |  |  |  |  |  | 325 |
| abdominal |  | 90 |  |  |  |  | 90 |
| mastectomy |  |  | 450 |  |  |  | 450 |
| hernia |  |  |  |  | 182 |  | 182 |
| knee |  | 71 |  | 84 |  | 148 | 303 |

**Supplementary Table 5**. *Summary of genome-wide significant hits, at SNP-,gene- and pathway-levels*. Colored sections are: Pink, overall summary indicating number of SNPs reaching P≤5×10^8^- (GWS), number of SNPs reaching P≤10^−7^ (NOMS), and number of genetic loci associated at GWS level (LOCI); Yellow, GWAS summary results from PLINK or METAL indicating the marker (SNP), the effect allele (A1) and alternative allele (A2), the effect allele frequency (A1FRQ), the marker’s chromosome (CHR) and base pair position (BP), the number of non-missing genotyped individuals (NMISS), the association effect size (BETA), standard error (SE) and P-value (P); Green, significant SNPs organized in genetic loci (LOCUS), locus’ chromosome (CHR) and approximate position (BP), with neighboring genes (GENES); Blue, GWAS summary results at gene-level from MAGMA indicating the gene (HUGO), number of aggregated SNPs (NSNPS) and number of genetic eigenvalues (NPARAM), the association test statistic (ZSTAT) and P-value (P_MULTI) corrected for genome-wide multiple testing with FDR (FDR_MULTI), from mean genetic effects’ P-value (P_SNPWISE_MEAN) and FDR-corrected (FDR_SNPWISE_MEAN), and from top one genetic effect’s P-value (P_SNPWISE_TOP1) FDR-corrected (FDR_SNPWISE_TOP1), the gene locus’ chromosome (CHR), start (START) and stop (STOP) position as well as the strand (STRAND), with up to ten most strongly associated SNPs in the gene (LEADING_EDGE_SNPS); Purple, GWAS summary results from MAGMA at pathway-level, indicating Gene Ontology’s pathway identifier (VARIABLE), the type of association (TYPE) which for pathways is ‘SET’, the number of genes in the pathway (NGENES), the association’s effect size (BETA), standard deviation (BETA_STD), standard error (SE), P-value (P), and FDR-corrected P-value (FDR), a short description of the pathway (DESC), and the top ten most strongly associated genes (LEADING_EDGE_GENES). Surgery types are: **(A)** hysterectomy; **(B)** abdominal; **(C)** mastectomy; **(D)** hernia; **(E)** knee; **(F)** meta-analysis of all the cohorts.

**Supplementary Table 6**. *Partitioned heritability in selected tissues and cell types*. Columns are: name of tissue or cell type, Name; the class of the tissue or cell type, Class; partitioned heritability coefficient (positive for enrichment), Coefficient; its standard error, Coefficient_std_error; its P-value, Coefficient_P_value; and the FDR-corrected P-value, FDR. Genes databases for partitioned heritability are from dataset published by: **(A)** Benita et al., **(B)** Fehrmann et al., **(C)** Trynka et al. For Benita, all results are shown, while those at the FDR 20% level are highlighted. For replication cohorts by Fehrmann and Trynka, only results at P<0.05 are shown. Column TableS1_Group refers to the group name assigned in Table S1 by Benita.

**Supplementary Figure 1.**
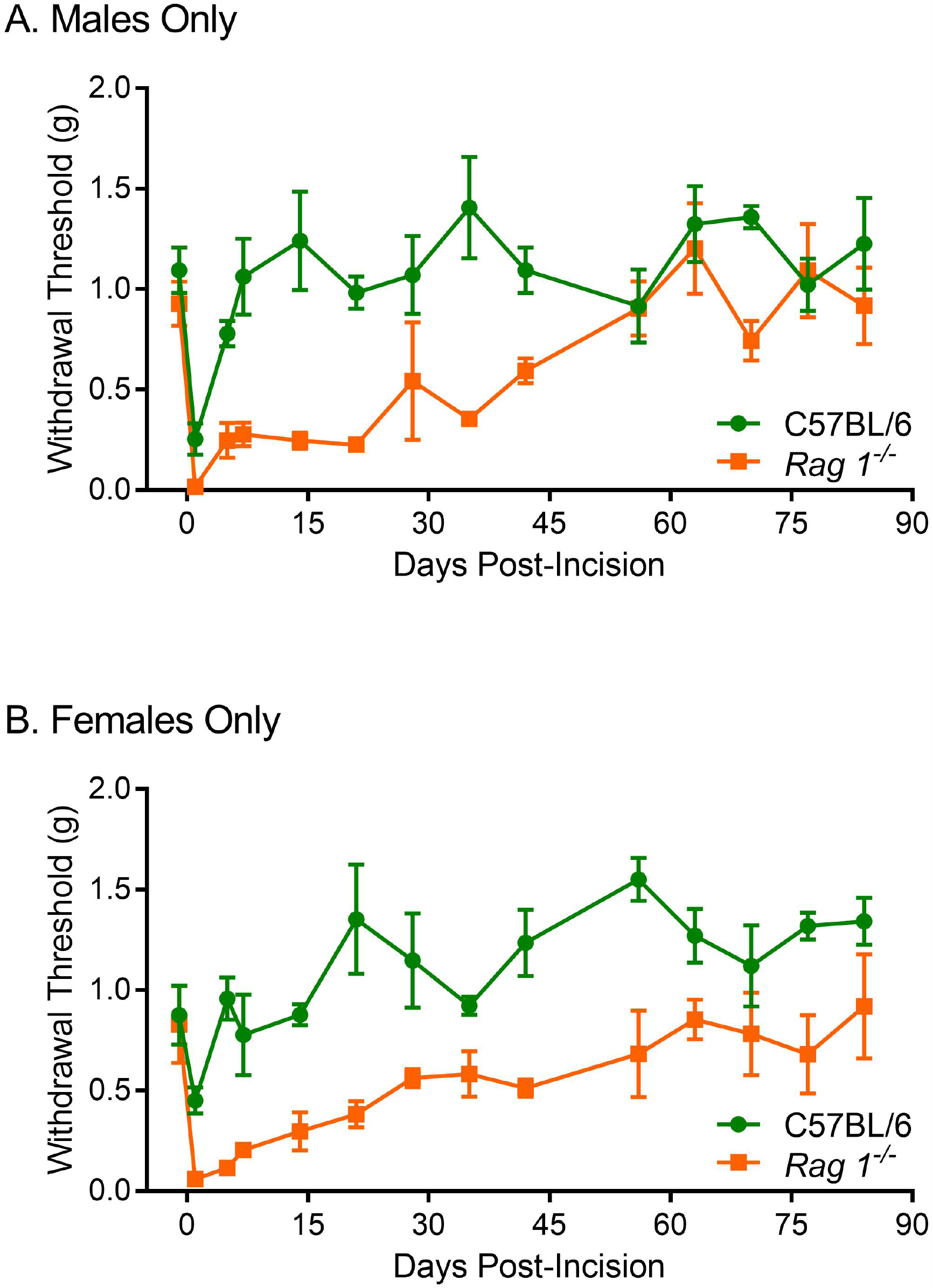
Sex-disaggregated plantar incision data. Symbols represent mean ± SEM hind-paw withdrawal thresholds (g) in male mice only (**A**; *n*=5 per genotype) and female mice only (**B**; *n*=5 per genotype).

